# National Implementation Trial of BeUpstanding™: An Online Initiative for Workers to Sit Less and Move More

**DOI:** 10.1101/2024.07.04.24309963

**Authors:** Genevieve N. Healy, Ana D. Goode, Lisa Ulyate, Alison Abbott, David Dunstan, Elizabeth G. Eakin, Nicholas D. Gilson, Lynn Gunning, Jodie Jetann, Anthony D. LaMontagne, Marj Moodie, Samantha Mulcahy, Neville Owen, Trevor Shilton, Leanne Sweeny, Leon Straker, Elisabeth A.H. Winkler

## Abstract

**Background:** The online BeUpstanding^TM^ program is an eight-week workplace-delivered intervention for desk-based workers to raise awareness of the benefits of sitting less and moving more and build a supportive culture for change. A workplace representative (the “champion”) delivers the program, which includes a workshop where teams collectively choose their sit less/move more strategies. A toolkit provides the champion with a step-by-step guide and associated resources to support program uptake, delivery, and evaluation. Here we report on the main findings from the Australian national implementation trial of BeUpstanding.

**Methods:** Recruitment (12/06/2019 to 30/09/2021) was supported by five policy and practice partners, with desk-based work teams from across Australia targeted. Effectiveness was measured via a single arm, repeated-measures trial. Data were collected via online surveys, toolkit analytics, and telephone calls with champions. The RE-AIM framework guided evaluation, with adoption/reach (number and characteristics); effectiveness (primary: self-reported workplace sitting time); implementation (completion of core components; costs); and, maintenance intentions reported here. Linear mixed models, correcting for cluster, were used for effectiveness, with reach, adoption, implementation, and maintenance outcomes described.

**Results:** Of the 1640 website users who signed-up to BeUpstanding during the recruitment period, 233 were eligible, 198 (85%) provided preliminary consent, and 118 (50.6%) champions consented and started the trial, with 94% (n=111 champions) completing. Trial participation was from across Australia and across industries, and reached 2,761 staff, with 2,248 participating in the staff survey(s): 65% female, 64% university educated; 16.9% from non-English speaking background. The program effectively changed workplace sitting (−38.5 [95%CI −46.0 to −28.7] minutes/8-hour workday) and all outcomes targeted by BeUpstanding (behaviours and culture), with small-to-moderate statistically-significant effects observed. All participating teams (n=94) completed at least 5/7 core steps; 72.4% completed all seven. Most champions spent $0 (72%) or >$0-$5 (10%) per team member; most (67/70 96%) intended to continue or repeat the program.

**Conclusions:** BeUpstanding can be adopted and successfully implemented by a range of workplaces, reach a diversity of staff, and be effective at creating a supportive culture for teams of desk-based workers to sit less and move more. Learnings will inform optimisation of the program for longer-term sustainability.

**Trial registration:** ACTRN12617000682347.

The trial was prospectively registered on the 12^th^ May, 2017 (ACTRN12617000682347), prior to the soft launch of the program, with the last update on the 11^th^ June, 2019 prior to the commencement of recruitment to the trial on the 12^th^ June, 2019.

## Introduction

High levels of sedentary time can be associated with poor health, wellbeing and productivity outcomes and indicators (1) with detrimental impacts particularly pronounced in those who are also physically inactive.(2, 3) The desk-based workplace has been identified as a key target setting for intervening on prolonged sedentary time,(4) with systematic review evidence concluding that interventions addressing prolonged workplace sedentary time can be both effective and acceptable.(5–9) Recent economic analyses have also indicated that reductions in sedentary time, to the levels achieved through workplace interventions, could result in Australian healthcare cost savings of $39 million per year.(10) Given this evidence, and the associated identification of workplace sedentary time as an emergent workplace health and safety issue,(11) there is demand from occupational policy and practice partners for an evidence-based, low-cost/no-cost, scalable solution to support workers and organisations to reduce prolonged sedentary time.(12)

The online BeUpstanding™ program (13) is based on a strong foundation of research evidence, including from randomised controlled trials (14) and best-practice approaches to workplace health promotion.(15) The 8-week program, targeted at desk-based work teams and using a champion-led approach, has a theoretical basis in social cognitive theory (16) and a social-ecologic model.(17, 18) The program is designed to raise awareness of the benefits of sitting less and moving more and create a culture of sustainable change through teams collectively identifying and promoting strategies to support these behaviours that are suitable for their work team and context.(19)

BeUpstanding has been iteratively developed and tested across multiple phases in collaboration with end users and key policy and practice stakeholders,(12, 20–23) and guided by the RE-AIM (reach, effectiveness, adoption, implementation, maintenance) framework.(24) Use of this framework is relevant for informing broader dissemination as it guides design and evaluation of programs in applied contexts.(25) This study reports on the adoption, reach, effectiveness, selected implementation, and maintenance intent outcomes from the Australian national implementation trial (Phase 4) of BeUpstanding.

Cost-benefit analyses, and detailed implementation, maintenance, engagement and process analyses, will be reported separately. Importantly, the implementation trial, which started in mid-June 2019, occurred in the context of major events in Australia and globally, namely the Australian Black Summer bushfires (July 2019 to March 2020) and the COVID-19 pandemic (March 2020--), which caused major disruptions to ways of working.(26, 27) Findings from this evaluation are intended to inform further optimisation of the BeUpstanding program prior to Phase 5 (dissemination).

## Methods

### Study design

Detailed methods for this implementation trial have been previously reported.(19) In brief, a single-arm design was used for evaluation of effectiveness, with repeated cross-sectional evaluations at pre-program (0 weeks), end-of-program (≈8 weeks; primary endpoint), and at 9 months post-program (≈12 months post sign-up). Only the pre-program and end-of-program data are reported herein, with data reporting according to the TIDieR (28) and TREND (29) checklists (Additional Files 1 and 2). The implementation trial was funded by a National Health and Medical Research Council (NHMRC) of Australia Partnership Project Grant (#1149936), which included cash and/or in-kind support from the five partners (Safe Work Australia, Comcare, Queensland Office of Industrial Relations, The Victorian Health Promotion Foundation (VicHealth), and Healthier Workplace Western Australia). These organisations are responsible for developing, implementing and/or promoting workplace health, safety and wellbeing in Australian workplaces. Ethical approval was from The University of Queensland Human Research Ethics Committee (approval #2016001743). Recruitment into the trial ceased on the 30^th^ September, 2021, noting the online program remained available for anyone to access and use. Only data from users who signed-up during the recruitment period are reported here.

### Participants and recruitment

Promotional efforts, co-ordinated by a detailed marketing and promotional plan developed by the research team and external marketing and communication experts, were made by the five partners to direct potential users to the BeUpstanding website (13) and the implementation trial. Multiple promotional channels were used, including social media, web links, email listservers, newsletters, workplace health promotion and occupational health networks, conferences, and workshops. Recruitment efforts were aimed at champions and decision makers of desk-based employees from a wide cross-section of industries, especially including the five sectors identified as priorities by the partners (regional, call centre, small business, blue-collar, and government).

Recruitment into the trial was a multi-step process. The BeUpstanding online program was freely available online for the entire trial period and beyond, without any requirement to be eligible or participate in the trial. A user could register (by completing preliminary details of themselves and their organisation) and fully sign-up to access the toolkit and associated resources by completing a champion profile survey and providing informed consent. Preliminary eligibility for the trial was ascertained from these two data sources (initial registration; champion profile survey), with eligibility being: not having previously run the BeUpstanding program; team size of at least five staff; and, most of the team do predominantly desk-based work. Those potentially eligible were contacted by telephone (call 1) by the research team to confirm criteria and ascertain the final eligibility requirements. Namely, that a workplace representative/employee signing up to the toolkit could perform the duties of a workplace champion and planned to run the program within the recruitment window. Potential champions were made aware that the main feature of taking part in the trial (as distinct from using the program independently) was they were committing to the evaluation steps in the BeUpstanding program and would receive expert health coaching support to deliver the program via email and phone calls.

All confirmed eligible champion participants were invited to participate in the trial, except where recruitment quota had been reached for a particular priority sector (see sample size) or a limit was reached for coaching availability to be allocated to a single organisation. Consent for the trial was in addition to that provided as part of the website sign-up process. The program was intended to be delivered to all staff in participating teams, with no exclusion criteria at the staff level. Staff provided informed consent as a step for gaining entry to the staff surveys.

### BeUpstanding program – national implementation trial version

A detailed description of the BeUpstanding program is reported elsewhere.(19) In brief, the program was designed to be run by the workplace champion across three phases (plan, do, review), each with associated tasks for the champion to complete. The accompanying toolkit provided information and training for the champion on the purpose of each phase and associated resources to support implementation, with the seven core components highlighted (starred) to indicate their importance. The most critical was the staff information and consultation workshop (or equivalent), where staff were provided information about the benefits of sitting less/moving more and collectively chose three (or more) team strategies they would use to sit less/move more as a team. Champions were encouraged to promote these strategies over eight weeks via the posters and emails provided in the toolkit. This participative approach meant each intervention program was unique for each team. In line with public, occupational, and clinical guidelines,(30–32) behavioural targets were for workers to achieve a 50:50 split between sitting and upright activities during work hours and to alternate sitting/upright posture at least every 30 minutes. Increased incidental movement throughout the day was also encouraged through the move more messaging. No incentives were provided as part of the trial. The main distinguishing feature of the implementation trial (as compared with the program in general) is that the research team both supported (and collected further data on) program implementation, via email support and telephone calls at five key timepoints: 1) recruitment; 2) obtaining consent; 3) at program initiation, following the staff consultation; 4) at end of program; and, 5) after approximately 12 months (maintenance). Project staff providing this support all had a minimal Masters level qualification and were experienced in motivational interviewing for behaviour change.

Due to the major shift to hybrid work resulting from COVID-19,(33) the toolkit and resources were audited and modified, and new resources were developed (e.g., see Additional File 3) during the trial to ensure the program was suitable for delivery for all desk-workers, no matter where they were working. These new resources went online in July 2020. Additional measures to capture work-from-home arrangements (staff completed) and the impact of COVID-19 on the workplace (champion completed) were also added (May 2020).

### Data collection

Detailed description of the data collection process is provided in the protocol paper.(19) Data were primarily collected via the dedicated, stand-alone BeUpstanding website, using surveys and toolkit analytics, with the project management team collecting and confirming implementation data via the telephone check-ins. Each toolkit user was required to complete the registration survey and champion profile survey as part of the sign-up process. Each champion was requested to complete a workplace audit before delivering the program and then a program completion survey after program delivery. Champions were also responsible for sending their bespoke links for the staff surveys (pre-program, post-program) to all staff in their team. For each survey, an anonymous staff identifier was constructed based on responses to three questions (month of birth; first initial of mothers first name; and, last three digits of mobile phone). This identifier, coupled with the cluster identification number, was used to ensure, per combined work team (cluster), each staff member responded only once and to match pre- and post-program responses. There was no blinding.

### Outcomes and Measures

The outcomes according to the RE-AIM framework are outlined below.

#### Adoption

Adoption was chiefly described in terms of uptake of the toolkit (n and % unlocking the toolkit), trial participation (n and %), the organisational, workplace, team and champion characteristics of those who participated in the trial, and how they had heard of BeUpstanding (in order to adopt it). Organisational characteristics were reported by champions, reconciled across champions from the same organisation, and checked against public records. Workplace characteristics were permitted to diverge between different champions from the same workplace. Champions were classified by project staff according to their role delivering the program to a team (overseer and primary champion / primary champion / co-champion) or supporting other aspects of BeUpstanding (overseer only / decision maker / other role). Further detailed information relevant to understanding adoption were the predictors of non-adoption (comparison of participants with non-participants), reasons given for not participating or withdrawing, and reasons for adopting the program, focusing specifically on what the champions said that they aimed to achieve by running BeUpstanding (open ended).

#### Reach

Reach outcomes included: the number of staff in participating teams (as estimated by the champion, and as verified through staff surveys), and the characteristics of staff who took part in the evaluation (by responding to one or both staff surveys).

#### Implementation

Implementation outcomes reported are completion rates (yes/no) of the seven core program steps, and champion-reported costs they incurred (reported in the program completion survey) or expected to incur (reported in the planning stage implementation check, shortly after selecting team strategies) through running BeUpstanding.

#### Effectiveness

Program effectiveness was assessed by the difference between pre-and post-program staff survey responses regarding 13 indicators of the program’s impact on workplace behaviour and workplace culture targeted directly by the program and 13 measures of productivity and health and wellbeing that were expected might improve as a result of the program. The primary outcome for effectiveness was workplace sitting, measured by the Occupational Sitting and Physical Activity Questionnaire (OSPAQ).(34) The post-staff survey also captured staff perceptions of the program and its impact, and adverse events. When described, adverse events were classified by two researchers as to whether they constituted adverse events and whether they could reasonably have resulted from the program. Program satisfaction was collected from staff in post-program surveys and from team champions in program completion surveys.

#### Maintenance intentions

The maintenance intentions of the champions of completing teams were ascertained by coding and reviewing the responses to an open text question of “What are your plans now in relation to BeUpstanding?” asked during program completion calls. Their detailed actual maintenance of the program and longer-term outcomes will be addressed in detail in subsequent papers.

### Sample size (primary effectiveness outcome)

As described elsewhere,(19) using assumptions informed by earlier iterations of the program (SD 90, r=0.5, ICC=0.1, post-attrition n/workplace = 5), it was determined that 47 to 62 workplaces (235 to 310 staff) were required to provide 80% to 90% power to detect a change in workplace sitting of at least 20 minutes per 8 hours of work (i.e., 4% of the workday) with 5% two-tailed significance. Consequently, the target sample size was at least 50 workplaces per priority sector to permit subgroup analysis.

### Statistical Analysis

Most of the reach, adoption, implementation and maintenance outcomes are described using descriptive statistics. Additionally, logistic regression was used to compare the organisational, workplace, team, and personal characteristics of those who included in the trial (trial adopters) versus non-participants (non-adopters), as well as those who completed the post-program evaluation of their team versus those who did not. Effectiveness outcomes (continuous) were compared between pre- and post-program staff surveys using linear mixed models, correcting for cluster (workplace) as a random intercept, or as random slopes, where likelihood ratio tests indicated the random slopes model was superior. Unadjusted models were reported in addition to the main findings, which adjusted for potential confounding due to different staff and teams providing data at each survey. Specifically, these were characteristics with a p<0.2 association either with teams being part of the post-program evaluation or not (tested by logistic regression models), or with staff members providing outcome data at only the pre-program survey (n=1481) versus only the post-program survey (n=397) (tested by mixed logistic regression models, correcting for workplace cluster as a random intercept). To test the sensitivity of conclusions to missing data handling, confounder-adjusted models were also reported with missing data imputed, using multiple imputation by chained equations. In addition to the analytic variables, imputation models contained auxiliary variables (additional variables predicting outcomes at either pre-or post-program surveys at p<0.2 in backwards eliminations) to help improve prediction of missing outcomes. A further sensitivity analysis excluded staff survey responses that had been collected after the program had already started. Analyses were performed in STATA version 18, with the final date of data extraction 15 May 2024. Significance was set at p<0.05 (two-tailed).

## Results

For ease of reporting, RE-AIM outcomes are presented these in the order of Adoption, Reach, Implementation, Effectiveness, and Maintenance intentions.

**Figure 1:**
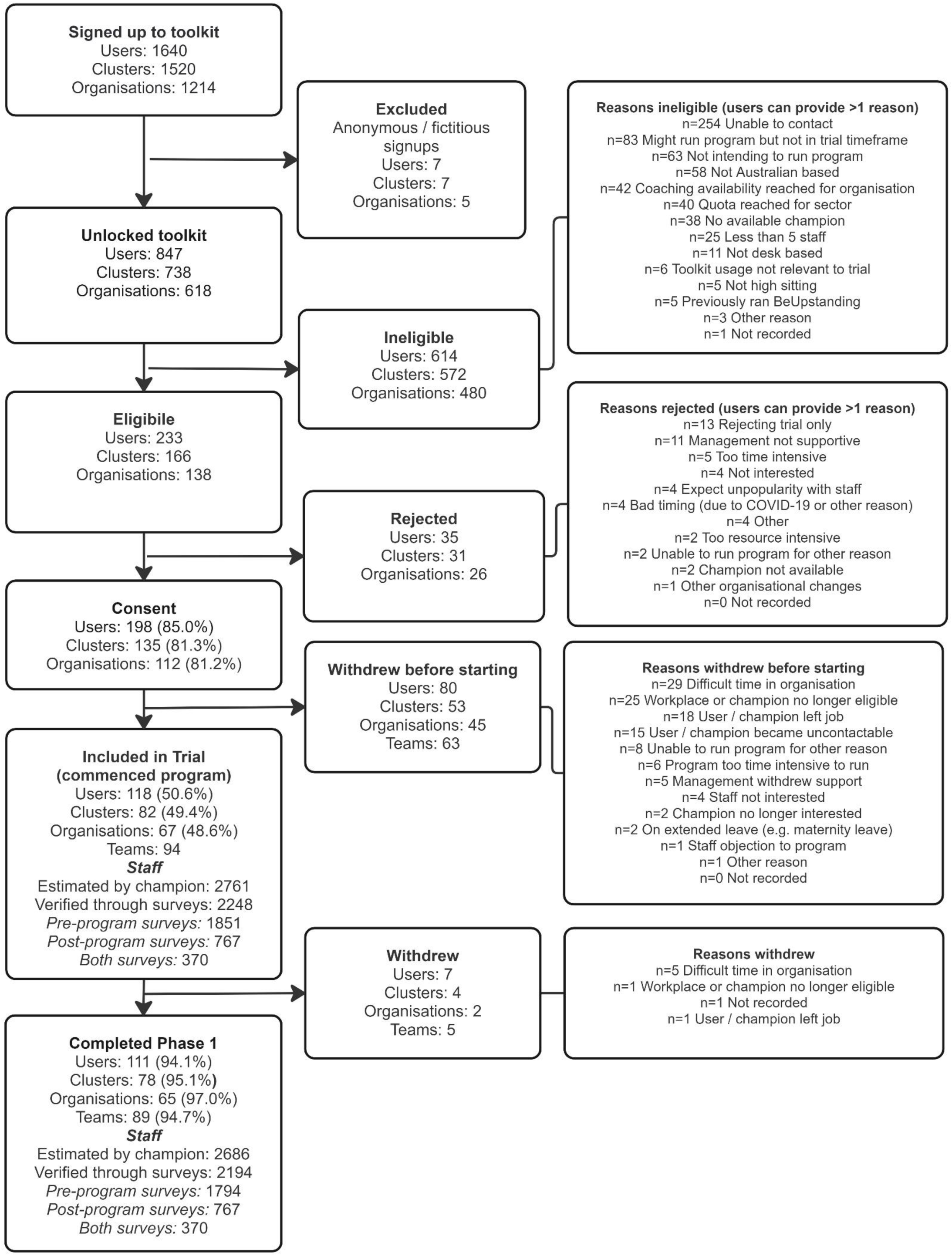
Participant flow diagram.

### Adoption

The participant flow diagram (Figure 1) shows that out of the 1640 website users who signed up during the trial recruitment period, 847 fully registered and unlocked the toolkit, and 233 were determined to be eligible for the trial, with ineligibility mostly due to not being contactable or not planning to run the program either at all or in the requisite timeframe. In total, 82 potentially would have been eligible if the recruitment quota for their sector(s) was considered not already met. Out of the 233 eligible users, the preliminary consent rate was 85.0% (n=198), which dropped to 50.6% (n=118) after excluding those who preliminarily seemed eligible and consented but withdrew before starting the program. Completion rates for the 118 who were included in the trial (who commenced the program) were high (94.1%, n=111). The most common reasons for non-participation prior to preliminary consent were reasons related to not being able to run the program, or lack of interest in the trial. Reasons for not participating after preliminarily consenting were primarily due to changing circumstances at either the organisation or champion level. Withdrawing after having started the program was rare and most often due to it being a difficult time in the organisation (n=5/7).

Almost all trial respondents (n=116) indicated what they aimed to achieve by adopting the program, with results categorised under 22 themes (Additional File 4). The most common aim was to improve either physical health and/or wellbeing (50.9%), with changing sitting / physical activity levels (37.9%) also often mentioned. There was some evidence that aims were associated with trial participation (Additional File 5). Reporting an aim to improve culture (OR=1.95, 95%CI: 1.09, 3.48), connection (OR=1.85, 95%CI: 1.02, 3.35), and/or productivity (OR=2.78, 95%CI: 1.47, 5.24) were associated with significantly higher odds of participation. Reporting a general aim to improve health was associated with a significantly lower odds of participation (OR=0.30, 95%CI: 0.09, 0.98).

Table 1 shows the characteristics of teams participating in the trial and their champions. The sample size target was met in terms of total numbers but not in any of the subgroups, due in part to the COVID-19 pandemic. Teams were recruited from every priority sector: 48 teams (51.1%) from the public sector; 24 teams (25.5%) from small-medium enterprises (11 (11.75%) small business); 33 teams (35.1%) from blue-collar workplaces; 32 teams (34.0%) with regional staff; and, six teams with call-centre staff (6.4%). Team size was highly varied and averaged 29.4 (SD=32.1) members. Overall, 13 of the 19 standard industry classifications in Australia (35) had some representation in the trial. The most common industry of the organisations of participating teams was Health Care and Social Assistance (n=16, 17.0%) and Professional / Technical and Scientific Services (n=16, 17.0%). Teams were recruited from every state and territory of Australia and were predominantly located in Queensland (n=43, 45.7%). Champions’ work locations varied widely in socioeconomic status, with a mean (SD) state percentile of 67.6 (26.7) in terms of socioeconomic index of advantage and disadvantage. Champions had a mean (SD) age of 42.9 (11.0) years. Many were female (75.9%), in middle (38.8%) or senior (15.5%) management, had a health and safety role (65.5%), and workplace health promotion training (56.9%). Less than half (36.2%) had prior experience delivering workplace health promotion programs.

**Table 1:**
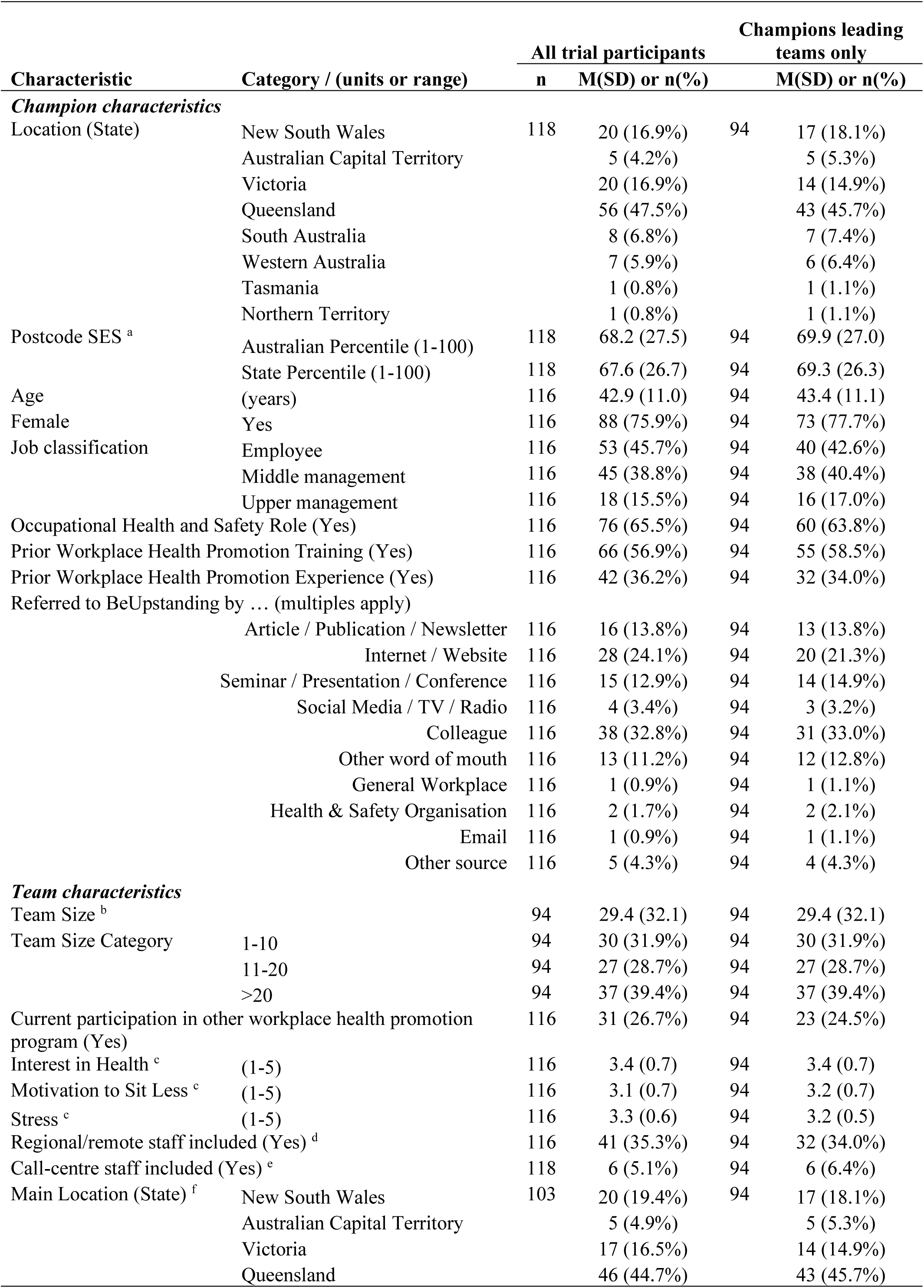
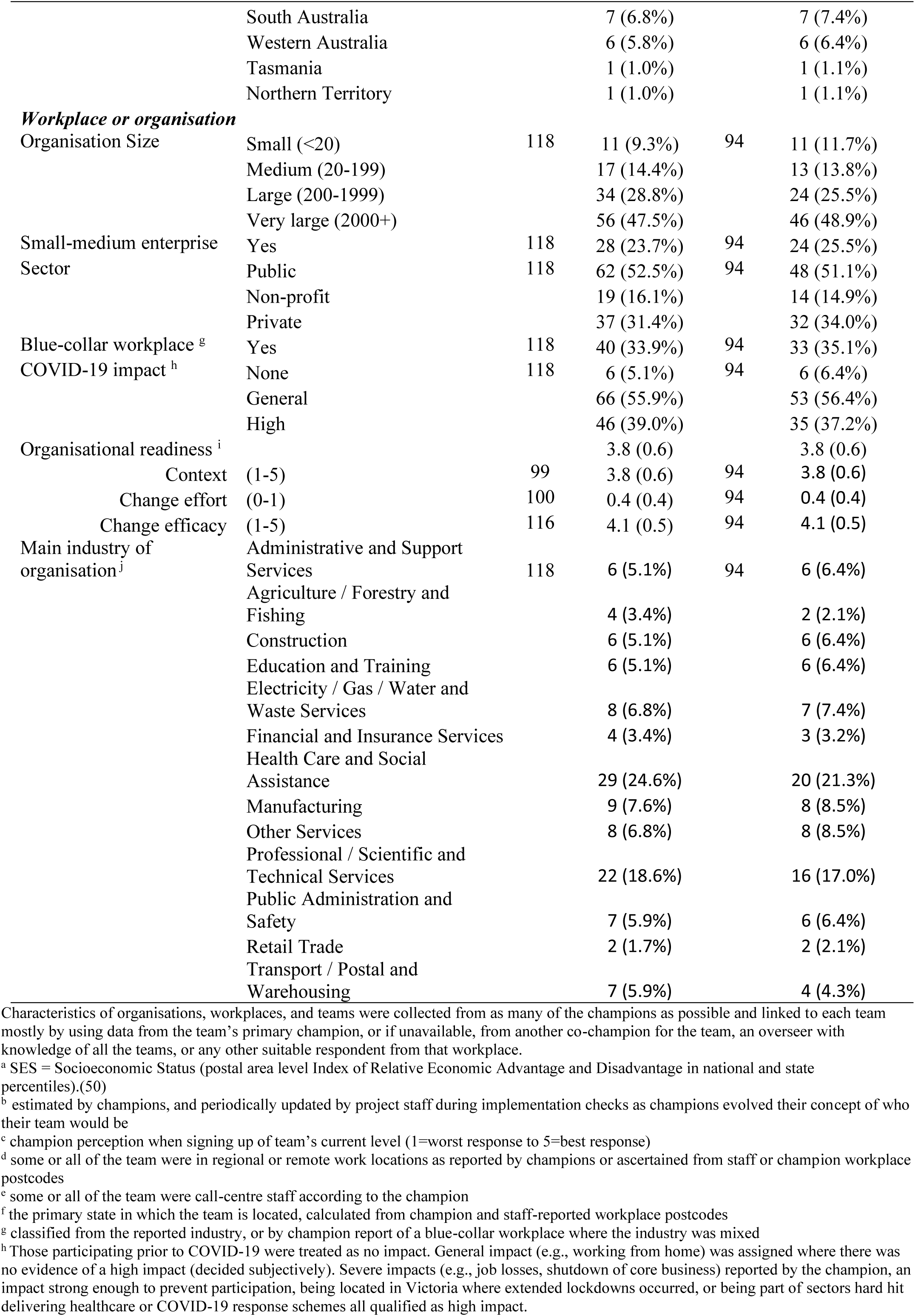
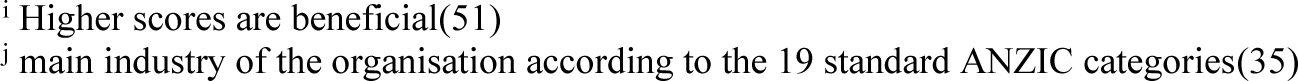
Characteristics of champions, teams and workplaces/organisations participating in the trial.

Additional File 5 shows the odds of participation (adopting the program) by these characteristics. Significant predictors included location (p<0.001, with highest participation in Queensland, and lowest participation in WA/NT/TAS), sector (p=0.001, with highest participation by public followed by non-profit then private sector), and COVID-19 impact level (p<0.001, with highest participation in high followed by general followed by no impact). Predictors significantly associated with lower odds of participation were: having an occupational health and safety role (0.60, 95% CI: 0.39, 0.91); inclusion of call centre staff (0.38, 95%CI: 0.16, 0.89); small-medium enterprise (0.50, 95%CI: 0.32, 0.79); and, hearing of the program through an article/publication/newsletter (0.56. 95% CI: 0.32, 0.98). Predictors significantly associated with higher odds of participation were: hearing of the program through a seminar/presentation/conference (1.97, 95%CI: 1.07, 3.64) or a colleague (2.29, 95%CI: 1.48, 3.52); and, higher organisational readiness (1.52, 95% CI: 1.09, 2.12 per unit change efficacy).

### Reach outcomes

Based on estimates from participating champions, the program reached 2761 staff in the trial, with 2248 able to be verified via responding to at least the identifier questions in one or both staff surveys. Staff were on average (mean (SD)) aged 42.9 (11.4) years, worked 38.2 (9.0) hours per week, and exercised for 3.1 (2.2) days per week prior to BeUpstanding, with 27.3% of staff meeting the recommended five or more days of exercise per week (Table 2). More than half of staff were female (65.0%), in full-time employment (82.1%), university educated (64.4%), non-managerial employees (68.5%), and in the highest occupational skill category (50.3%). A very small minority were shift-workers (2.1%), and 16.9% of staff were from a non-English speaking background.

**Table 2:** Characteristics of participating staff (n=2228)

| Characteristic | (Unit or range) / category | n | Mean (SD) or n(%)<sup>a</sup> |
| --- | --- | --- | --- |
| Age | (years) | 2004 | 42.9 (11.4) |
| Work hours | (hours per week) | 2043 | 38.2 (9.0) |
| Pre-program exercise <sup>b</sup> | (days per week) | 1768 | 3.1 (2.2) |
| Meeting recommendations <sup>b</sup> | No <5 days per week | 1768 | 1286 (72.7%) |
|  | Yes, ≥5 days per week |  | 482 (27.3%) |
| Post-program exercise <sup>b</sup> | (days per week) | 709 | 3.5 (2.1) |
| Meeting recommendations <sup>b</sup> | No, <5 days per week | 709 | 469 (66.1%) |
|  | Yes, ≥5 days per week |  | 240 (33.9%) |
| Body Mass Index (BMI) <sup>c</sup> | <25 kg/m<sup>2</sup> | 1705 | 771 (45.2%) |
|  | 25-<30 kg/m<sup>2</sup> |  | 561 (32.9%) |
|  | ≥30 kg/m<sup>2</sup> |  | 373 (21.9%) |
| Female | No | 2131 | 745 (35.0%) |
|  | Yes |  | 1386 (65.0%) |
| Non-English speaking background | No | 2015 | 1674 (83.1%) |
|  | Yes |  | 341 (16.9%) |
| Full-time employment | No | 2043 | 365 (17.9%) |
|  | Yes |  | 1678 (82.1%) |
| Education | High school or less | 2024 | 191 (9.4%) |
|  | TAFE/Trade certificate/<br>Diploma |  | 530 (26.2%) |
|  | University or other tertiary |  | 1303 (64.4%) |
| Job classification | Employee | 2043 | 1399 (68.5%) |
|  | Middle management |  | 167 (8.2%) |
|  | Upper management |  | 477 (23.3%) |
| Job category skill level <sup>d</sup> | 1 (most skill / training) | 2043 | 1028 (50.3%) |
|  | 2 |  | 266 (13.0%) |
|  | 3 |  | 505 (24.7%) |
|  | 4 |  | 56 (2.7%) |
|  | 5 (least skill / training) |  | 188 (9.2%) |
| Shift worker | No | 2043 | 2001 (97.9%) |
|  | Yes |  | 42 (2.1%) |

<sup>a</sup> Mean SD corrected for clustering using linearized variance (STATA survey commands)
<sup>b</sup> number of days per week exercised for at least 30 minutes (0-7)(52) and meeting national guidelines (≥5 v <5 days)
<sup>c</sup> Excludes invalid responses (truncated to plausible range of 15 to 50)(53)
<sup>d</sup> Australian Standard Classification of Occupations (ASCO: 1 most to 5 least skilled)(54)

### Implementation outcomes

The champions had completed all core steps for most of the participating teams (72.4%) and most of the completing teams (76.4%). The steps with the lowest completion rates were the workplace audit, strategy selection, and the program completion survey (Table 3). During their planning stage implementation check-in calls, shortly after selecting their team strategies, 80 champions reported on any costs they had incurred or expected to incur in order to support or enact their team strategies. In total, 35 cost items were reported, the most common of which was sit-stand desks or similar (n=14). Other costs included: accessories to assist with standing at the desk (Bluetooth headsets (n=3); anti-fatigue mats (n=1)), aids for prompting (reminder/lockout software (n=4); timer (n=1)), enhancements to communal areas (standing benches/tables (n=3); communal bins (n=1)), equipment (n=2) and subscriptions (n=1) for physical activity at work; and, miscellaneous other (pedometers (n=1); undescribed equipment (n=2); sporting goods gift cards (n=1)). In the program completion survey, 72 champions reported what was spent in total to run the program for their team, with the majority reporting spending $0 (n=52, 72.2%, Table 4). Sit-stand desks had been purchased for every team where the amount spent was >$1000.

**Table 3:** Completion of core steps for all participating teams (n=94) and completing teams (n=89) by one or more champion(s) for that team.

| Core program steps | All teams<br>n (%) | Completing teams<br>n (%) |
| --- | --- | --- |
| - Pre-program staff survey | 94 (100.0%) | 89 (100.0%) |
| - Workplace audit <sup>a</sup> | 71 (75.5%) | 71 (79.8%) |
| - Staff information and consultation session (e.g., workshop) | 94 (100.0%) | 89 (100.0%) |
| - Strategy selection | 73 (77.7%) | 78 (87.6%) |
| - Program promotion via posters and/or emails | 94 (100.0%) | 89 (100.0%) |
| - Post-program staff survey | 94 (100.0%) | 89 (100.0%) |
| - Program completion survey | 80 (85.1%) | 79 (88.8%) |
| Number of core steps completed (out of 7) |  |  |
| 7 | 68 (72.3%) | 68 (76.4%) |
| 6 | 17 (18.1%) | 16 (18.0%) |
| 5 | 9 (9.5%) | 5 (5.6%) |
| <5 | 0 (0.0%) | 0 (0.0%) |

<sup>a</sup> The workplace audit mostly asked about the workplace in general and was considered completed if it was completed by any champion for the workplace

**Table 4:**
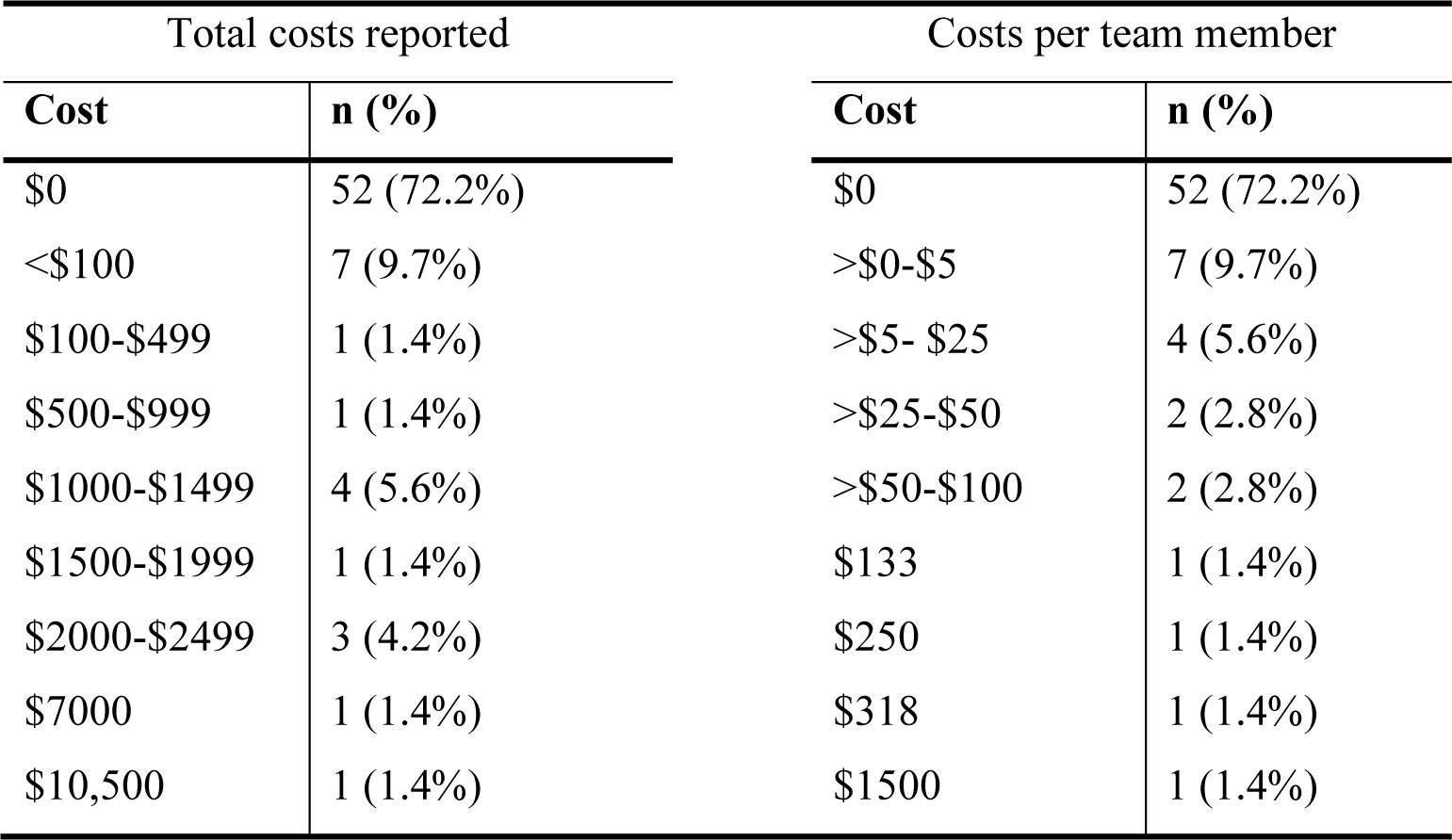
Post-program costs reported by the champion (n=72 champions)

| Total costs reported |  | Costs per team member |  |
| --- | --- | --- | --- |
| Cost | n (%) | Cost | n (%) |
| \$0 | 52 (72.2%) | \$0 | 52 (72.2%) |
| <\$100 | 7 (9.7%) | >\$0-\$5 | 7 (9.7%) |
| \$100-\$499 | 1 (1.4%) | >\$5- \$25 | 4 (5.6%) |
| \$500-\$999 | 1 (1.4%) | >\$25-\$50 | 2 (2.8%) |
| \$1000-\$1499 | 4 (5.6%) | >\$50-\$100 | 2 (2.8%) |
| \$1500-\$1999 | 1 (1.4%) | \$133 | 1 (1.4%) |
| \$2000-\$2499 | 3 (4.2%) | \$250 | 1 (1.4%) |
| \$7000 | 1 (1.4%) | \$318 | 1 (1.4%) |
| \$10,500 | 1 (1.4%) | \$1500 | 1 (1.4%) |

### Effectiveness outcomes

#### Outcomes targeted by the intervention

Baseline levels of the staff-reported program outcomes and their changes over the program are shown in Table 5. The main models adjusted for characteristics that might differ between those providing data at post- and pre-program surveys, based on participation in the post-program evaluation by the team (Additional File 6) or staff member (Additional File 7). These models (Figure 2) indicated significant improvements in all outcomes directly targeted by the program: the primary outcome (self-reported work sitting); work standing and moving; the extent to which sitting was accumulated in prolonged bouts; the alignment between preferred and actual sitting, standing and moving at work; and, all measures of workplace culture. The magnitude of these effects on average were mostly small-to-moderate, with the smallest effects seen for moving, moving alignment, and perceived control over sitting. Workplaces varied significantly in their changes in activity at work and sitting in prolonged bouts, with tests for random slopes reaching statistical significance (Additional File 8).

**Figure 2:**
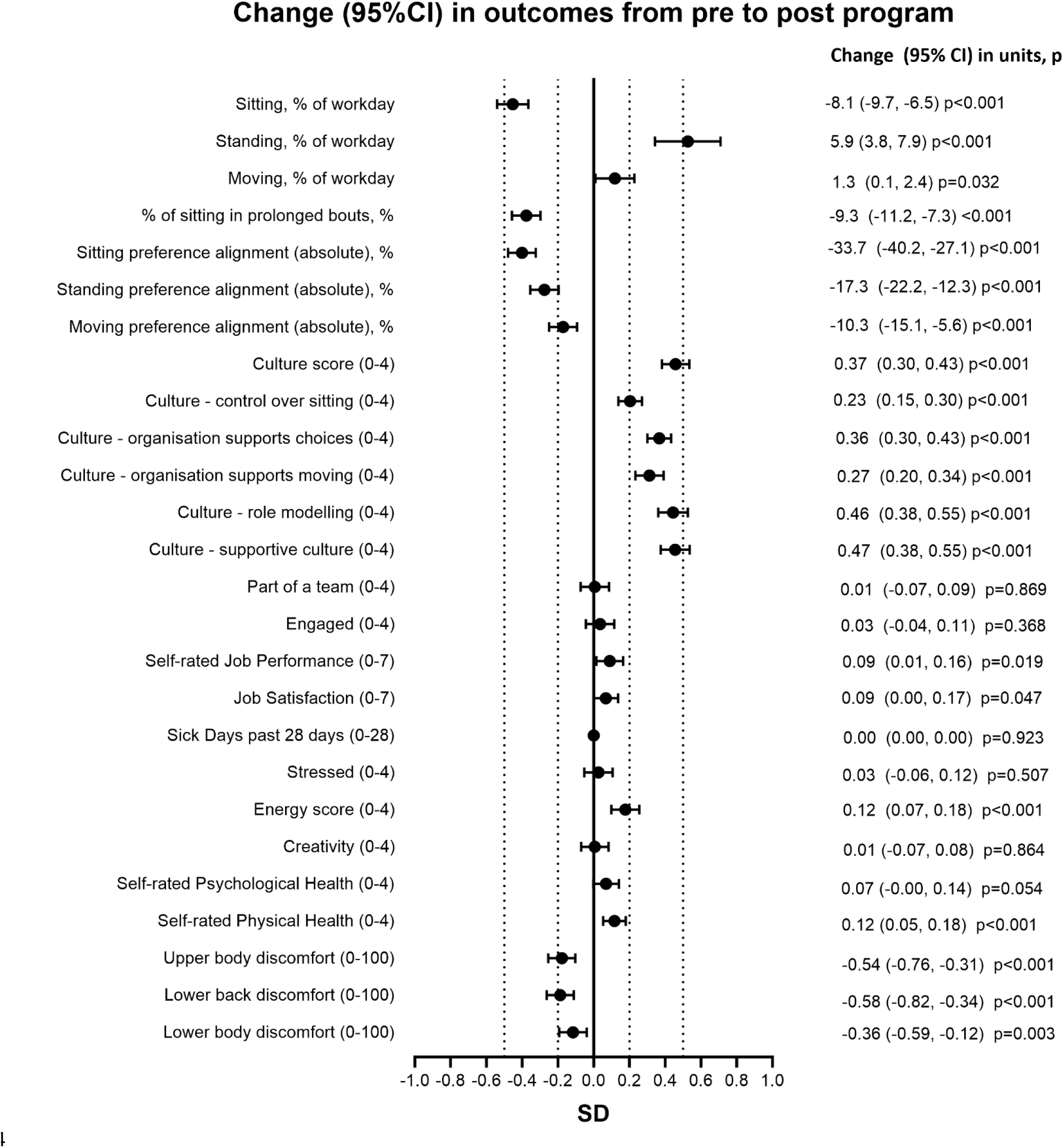
Change in staff-reported outcomes from pre to post BeUpstanding intervention.

**Table 5:**
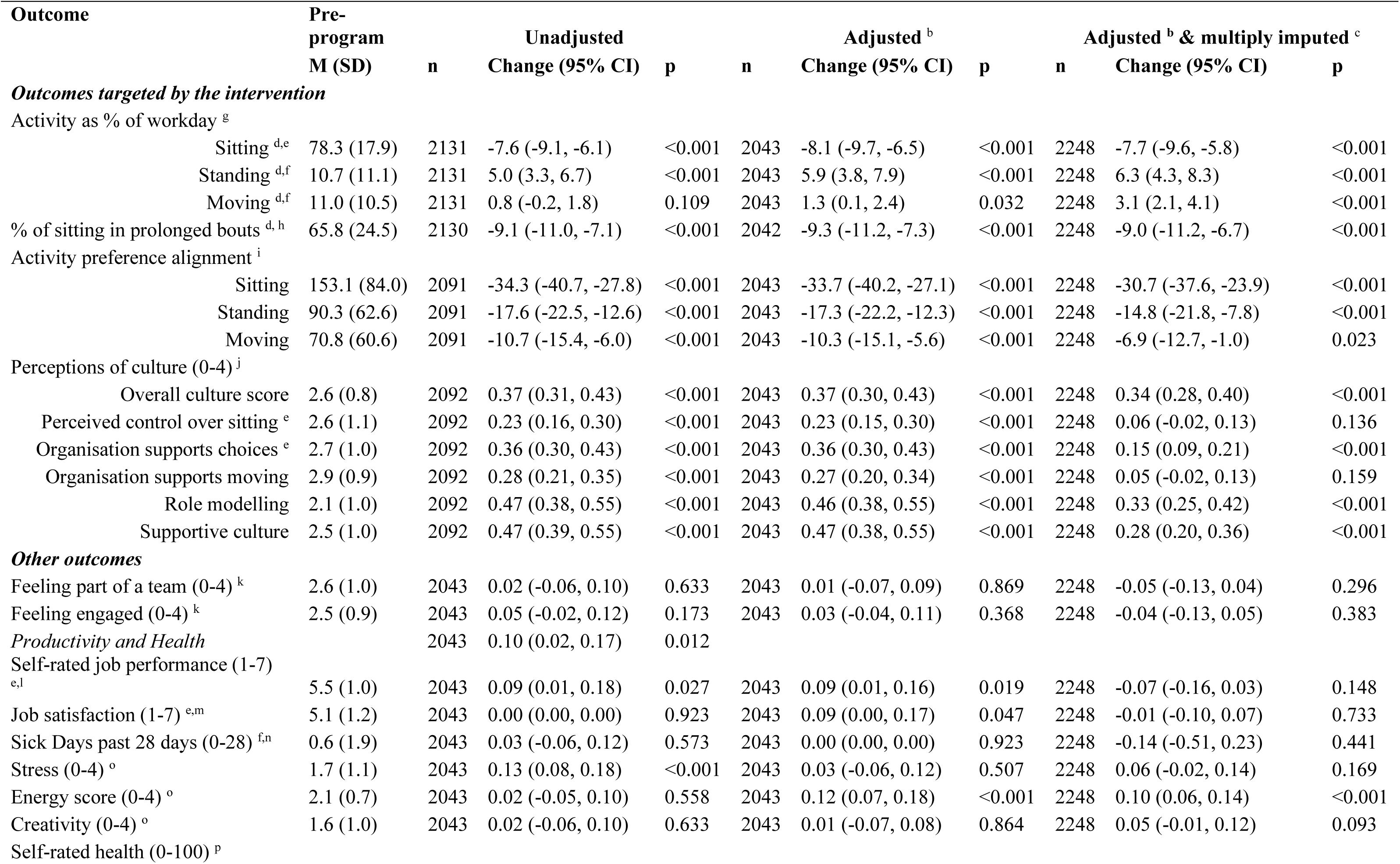
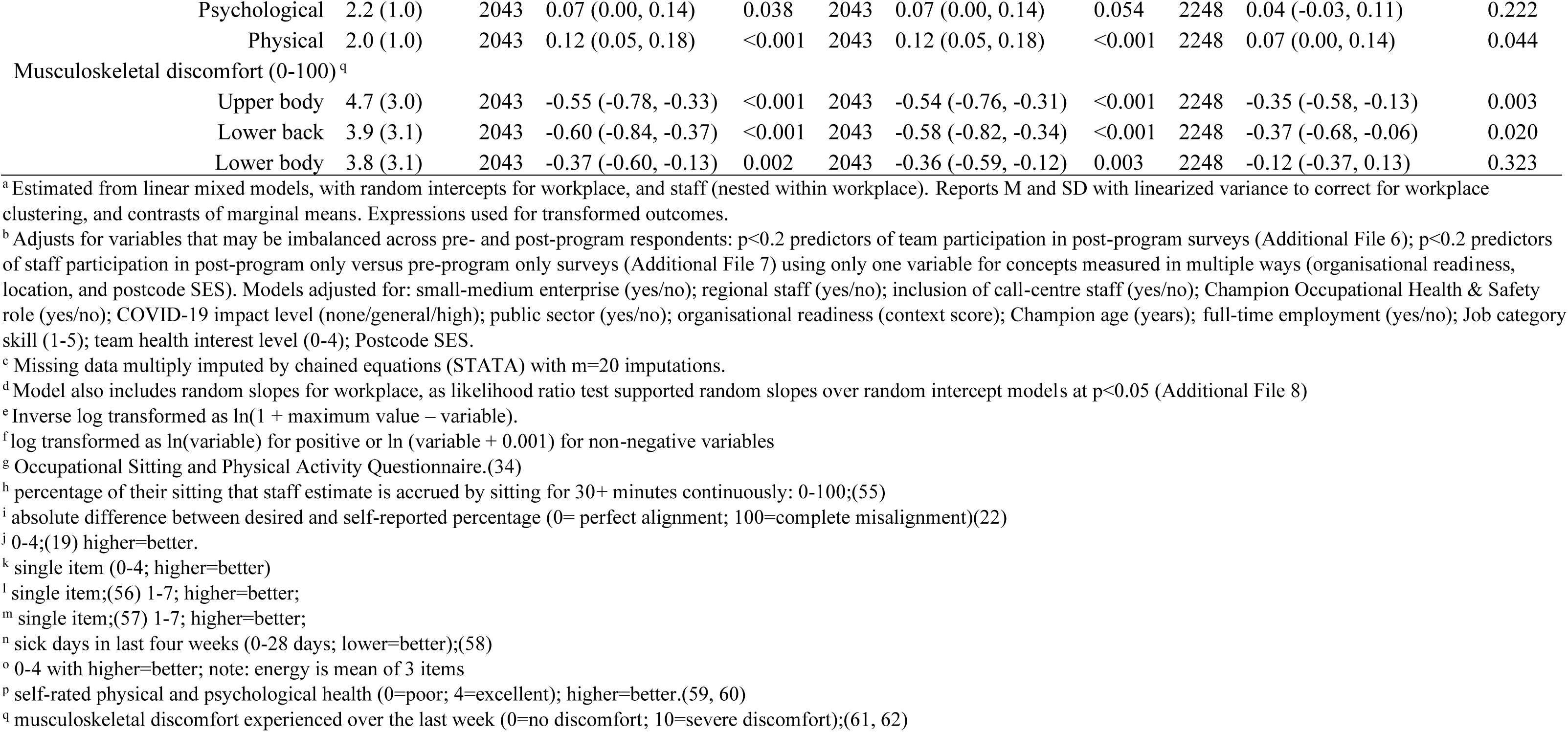
Changes in staff-reported main outcomes pre to post BeUpstanding program (k=82 workplaces) ^a^.

Conclusions regarding workplace behaviours from the main adjusted models were mostly robust to modelling choices, except that moving results strengthened slightly and became significant in adjusted models and then further strengthened with imputation. Results concerning culture were unaffected by statistical adjustment but were sensitive to missing data handling, whereas results from imputation models were often weaker and sometimes no longer statistically significant. Excluding the pre-program surveys that had been collected after the program commenced had no meaningful impact on findings (Additional File 9).

#### Other outcomes

Of the 13 outcomes concerning work productivity and health, significant improvements were seen in self-rated job performance, job satisfaction, energy, self-rated physical health, and musculoskeletal discomfort in the upper body, lower back, and lower body, with improvements ranging in magnitude from very small to small (Figure 2; Table 5). Other changes were consistently all very small, and not statistically significant. Random slopes tests were not statistically significant for any of these outcomes (Additional File 8). Findings were similar to those from unadjusted models, except that adjustment brought results for engagement even closer to the null and the previously significant change in self-rated psychological health became borderline significant. There were no meaningful changes in outcomes excluding late pre-program surveys (Additional File 9). Many findings, especially for work-related and productivity outcomes, were highly sensitive to missing data, where multiple imputation results only indicated significant changes (all improvements) in energy, self-rated physical health, and musculoskeletal discomfort in the upper body and lower back, all of which were somewhat smaller than they had been in the evaluable case analyses. In imputed models, effects for lower body discomfort were somewhat attenuated in magnitude and no longer statistically significant, while effects for estimates of changes in self-rated job performance and job satisfaction were completely attenuated. Effect sizes became further away from the null for sick days, stress, and creativity, but remained very small and did not reach statistical significance.

#### Priority sectors

The changes in the primary outcome (work sitting) overall amounted on average to −38.5 (−46.0, −31.0) min/8 h for the trial in general. Changes were mostly also present within each of the priority sectors: −28.2 (−39.7 to −16.6) min/8-h in small-medium enterprises (based on 24 workplaces and 496 staff); −35.8 (−49.4 to −22.1) min/8-h in blue-collar workplaces (based on 26 workplaces and 772 staff), −41.7 (−52.5 to −31.0) min/8-h in public-sector workplaces (based on 40 workplaces and 954 staff); and, −45.2 (−60.0 to −30.3) min/8-h in teams that included regional staff (based on 26 workplaces and 626 staff). Changes in the teams that included call-centre staff were −46.3 (−126.4 to 33.9) min/8-h and did not reach statistical significance with the small sample of 3 workplaces and 97 staff.

#### Adverse events and staff perceptions of the program

Most (n=604, 96.0%) of the 629 staff responding to the staff follow-up surveys did not experience any adverse events. Only 7 staff (1.1%) reported an adverse event that was confirmed by research staff, with six of these (1.0%) potentially being attributable to the study, and a further 18 staff (2.9%) reported that they experienced some adverse event that could not be verified as they did not describe it. The six events that could have arisen from program participation were: new or exacerbation of musculoskeletal pain or injury (n=4), specifically to the hip from standing propped on a stool, to the back from “overdoing things”, plantar fasciitis, and lower leg and lower back pain; and, adverse work impacts (n=2), specifically feeling guilty about leaving the desk (n=1) and feeling followed by their supervisor (n=1).

The majority of staff reported positive impacts on outcomes targeted directly by BeUpstanding (culture, knowledge, attitudes, awareness), with few reporting negative impacts (Figure 3). Staff provided more mixed responses in relation to the impact of the program on activity outside work, with very few staff (n=11, 1.7%) perceiving a negative impact and the rest of the staff being almost equally divided between perceiving a negligible impact (51.1%) or a positive impact (47.2%).

**Figure 3:**
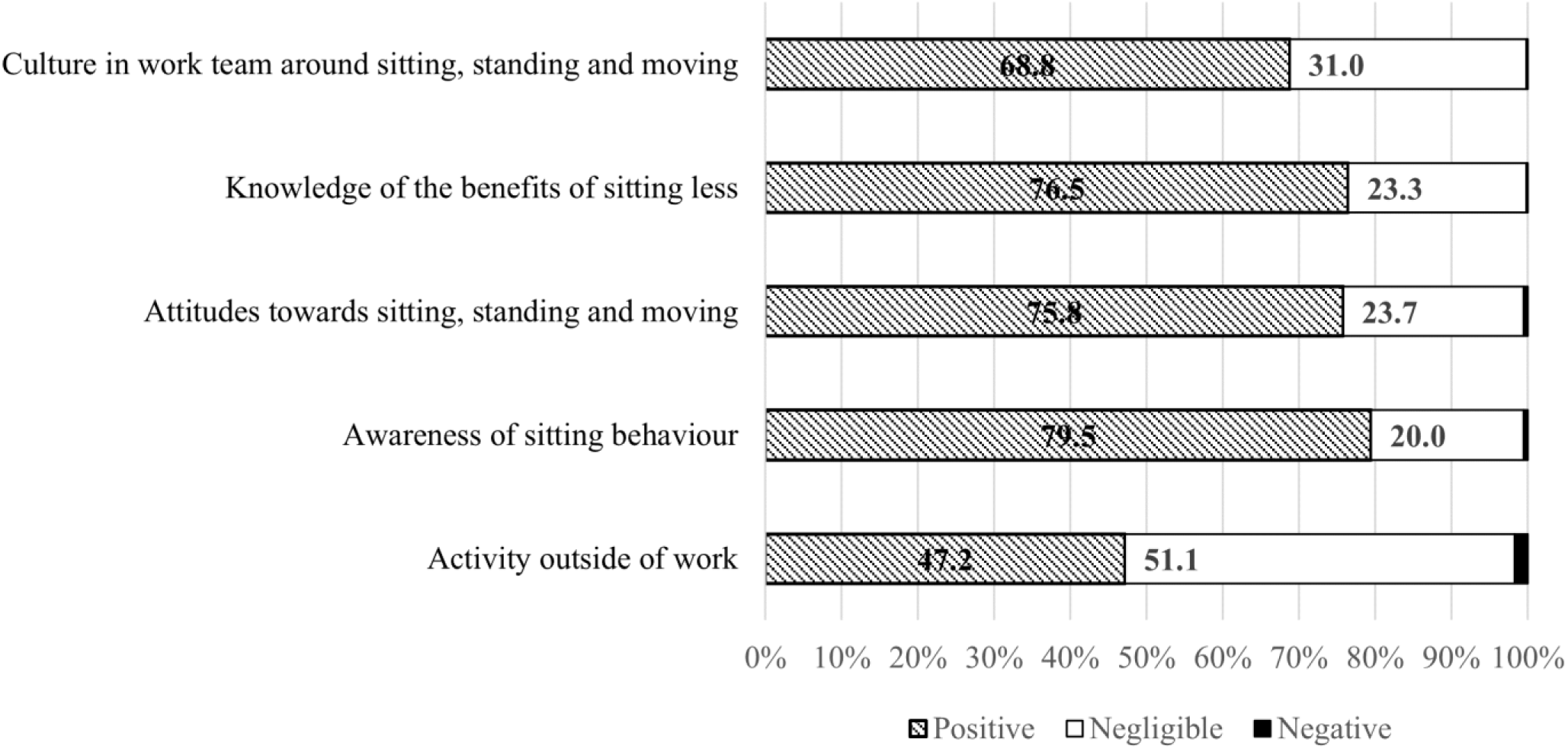
Staff perceptions of the impact of the BeUpstanding program (n=654)

#### Program satisfaction

Staff (n=654) reported on their satisfaction with the program. Excluding those who indicated they were not aware of BeUpstanding or responded not applicable (n=35), most staff (n=477, 77.1%) indicated they enjoyed BeUpstanding, some were uncertain (n=115, 18.6%), and a minority of staff said they did not enjoy participating (n=27, 4.4%). Champions also reported on their satisfaction with the program in post-completion surveys (n=72). When asked whether the program met expectations, most said yes (n=57, 79.2%) or somewhat (n=13, 18.1%) and only two (2.8%) said no. Similarly, most said they would (n=63, 87.5%) or maybe would (n=8, 11.1%) recommend the program and only one said they would not (1.4%).

#### Maintenance intentions

The 8-week BeUpstanding program was completed by 89 of the 94 teams that were part of the trial. Upon completion, project staff were able to contact 70 of their team champions and ask about their planned next steps concerning BeUpstanding. Most responses to the open-ended question indicated the champions intended to continue the program (n=59, 84.3%) or repeat it (n=8, 11.4%), while very few indicated that the champion intended to stop or pause the program (n=1, 1.4%), including to restart it with different staff (n=1, 1.4%), and one champion intended to check with management (n=1, 1.4%).

## Discussion

This study reports the main outcomes, covering the five dimensions of the RE-AIM framework, of the first national implementation trial of a workplace health promotion initiative specifically supporting teams of desk-based workers to sit less and move more (BeUpstanding). This trial was the fourth phase in the research-to-practice translation process from an intervention that had demonstrated effectiveness in the context of a cluster randomised controlled trial.(14)

### Adoption and Reach

While there was interest in the program more broadly, with over 600 organisations unlocking the toolkit during the trial period, adoption of the program as part of the research trial was more modest, with approximately half of eligible workplaces running the program. This needs to be understood in the context of the BeUpstanding program being available for free online without participation in the trial, and two significant emergencies in Australia that occurred during the trial’s recruitment period, specifically the COVID-19 pandemic and the 2019-20 bushfires. Reported reasons for not adopting the program were mostly regarding it being a difficult time in the organisation (e.g., availability of personnel) or lack of interest in the research trial. Only rarely did withdrawal occur after initial consent due to an issue with the program. As might be expected, a higher level of organisational readiness, specifically change efficacy, was significantly associated with higher odds of adoption into the trial while conversely having an occupational health and safety role was associated with lower odds of adoption. One interpretation (among many) is that champions who do not feel their workplace is well placed to run the program may require additional supports (beyond what was provided in the implementation trial) to adopt the program, while conversely those in occupational health and safety roles may have felt confident to run the program independently outside of the trial context without support. Other reasons could include that occupational health and safety staff had competing priorities especially in the COVID-19 pandemic, and unready workplaces may have wanted to avoid the oversight of a trial.

Workplaces and staff took part from every partner-identified priority sector, from every Australian state and territory (including rural/regional locations), from 13 of 19 ANZIC industries,(35) and with diverse workplace and worker characteristics. Some groups were easier to reach than others, with limited participation from shift workers, from those not in fulltime employment, and with lower-skilled occupations. While this may align with our specific focus on desk-based workers, it is important in future research to understand how this program and others like it may work in these groups, as well as other underrepresented or untested key populations, such as people with disabilities. Adoption was significantly lower for workplaces outside the public sector, of small-medium size, and especially in teams that included call-centre staff (despite deliberate recruitment efforts). Staff in call centres or contact centres generally have higher rates of sedentary time than general office workers,(36) but typically also have reduced autonomy to break up their sedentary time due to their job tasks and job demands.(37) In line with previous recommendations,(38) it will be important to work with relevant stakeholders to co-design appropriate approaches for this particular setting.

### Implementation

Champions were provided automated feedback via the toolkit that was engaging and meaningful and had a streamlined online user experience that highlighted the core components. As part of the trial, they also received support from the research team at key stages: all factors that had been added and optimised based on learnings from the version used by the early adopters.(21) In the previous phase of this research-to-practice translation (Phase 3), the seven core steps were completed for only 5% of teams, due in part to retention and in part to progressing through without completing core steps.(20) These modifications appear to have resulted in a much more complete implementation of the program, with most teams (>72%) completing all seven core steps, and none completing fewer than five. Most workplaces implemented the program in a no or low-cost manner, and only a minority spent significant sums of money, usually on sit-stand workstations.

### Effectiveness

The program was significantly effective in relation to the primary outcome (workplace sitting), behaviours and culture (which were the main targets of the program), and also for some indicators of productivity, health and wellbeing. The average decrease in self-reported work sitting of −38.5 (95%CI −46.0 to −31.0) minutes per 8-hour workday appeared to have co-occurred mainly with increased standing, and also a smaller amount of increased moving. Notably the extent of change strongly resembled the average effect on sedentary behaviour of 38 minutes reduction per workday (95% CI −47.3 to −28.7) achieved by sedentary behaviour interventions in office workers (across all intervention types combined) as reported by a recent systematic review and meta-analysis.(39) Given the timing of the trial, and the substantial shift to hybrid work resulting from the COVID-19 pandemic,(33) these findings also provide some evidence that BeUpstanding is suitable for work from home and hybrid work contexts. A pre-post design was required for feasibility reasons, leaving open the possibility of other explanations for changes over time, especially in light of the pandemic. Notably however, the controlled studies that informed the BeUpstanding program development had not observed any meaningful degree of changes in control groups.(14, 40, 41) It is not likely that the pandemic produced the improvements, considering COVID-19 saw an increase in sedentary time and a reduction in physical activity in desk workers(42), and the average change was −43 minutes per 8=hour workday (− 9.0%, 95% CI: − 12.0% to − 5.9%) in the pre-pandemic evaluation of the program’s early adopters.(20)

Findings concerning workplace sitting, other workplace behaviours and workplace culture were robust to issues of potentially selective evaluation, being largely unaltered in sensitivity analyses. Where selective participation may have been more of an issue was in gauging the impact of the program on productivity and health, where changes were typically smaller after accounting for missing data, but notably were still present for energy, self-rated physical health, and upper body and lower back musculoskeletal discomfort. The benefits seen to at least some of the productivity and health indicators adds to the growing evidence from field-based studies that interventions aimed at reducing workplace sedentary time, particularly those where sedentary time is predominantly replaced with standing, are not detrimental to indicators of work performance.(43) The musculoskeletal findings in particular add to the currently limited evidence base in this area.(17) Although these reductions were small on average, they are potentially important given that work-related musculoskeletal disorders are the leading work health and safety issue in Australia in terms of both frequency and costs,(44) and were not necessarily small for all participants. Future detailed evaluation will explore whether changes are possibly enhanced based on factors such as pre-existing discomfort, and nuanced aspects of the behaviour changes, such as the reallocation of sitting to standing versus to moving, the prolonged or interrupted nature of sedentary accumulation, and the specific behavioural strategies that were adopted.

A key goal of the implementation trial was to evaluate effectiveness in not just overall but also within key sectors identified *a priori.*(19) Here, significant improvements in workplace sitting were seen within almost all sectoral subgroups. The sole exception was that changes in teams with call centre staff were substantial −46.3 minutes per 8-hour workday (95%CI: −126.4 to 33.9) but inconclusive with wide confidence intervals as a consequence of only three call centres participating in the evaluation. This finding adds to a known paucity of evidence, as indicated by a recent scoping review(45) that called for further evidence on the effectiveness, acceptability and feasibility of health-promoting interventions (including for addressing sedentary behaviour) conducted in the call-centre setting. Minimal adverse events were recorded and were of mostly a similar nature (musculoskeletal) as have been reported in similar programs.(46, 47)

### Satisfaction and Maintenance intentions

Satisfaction was generally high for both staff and champions and most indicated they would participate again in BeUpstanding. Although further evidence is needed,(48) shifting culture may be the key for sustainable change in workplace sedentary behaviour. The BeUpstanding program heavily targeted culture, and significantly improved many cultural aspects. Following BeUpstanding, most champions intended to continue on with the program, often also intending adaptation --an important feature of sustainable delivery.(49) Many particularly mentioned intentions to expand to either more staff and/or embedding the program into standard practice, which is consistent with the intent of BeUpstanding. Findings from the 12-month follow-up champions (to be reported elsewhere), will provide further insights into workplace changes following BeUpstanding.

### Strengths and limitations

Strengths of the study include: the wide-ranging participation from workplaces across Australia and from a wide variety of industries and all priority sectors identified by the policy and practice partners; the data collection across the RE-AIM framework, enabling understanding of how the program is working in practice; and, the data collection on a range of measures, including those relevant to organisations (e.g., job satisfaction). Limitations include the single-group evaluation design, which was necessary for feasibility reasons, and the large workforce turnover, reflective of the broader job mobility occurring during this period.(33) To some extent this was mitigated by combining longitudinal and repeated cross-sectional designs, and using statistics that account for repeated observations but do not require them. Staff could be included in the evaluation whether they were part of the team and participating in the evaluation in either one or both evaluations. Another limitation was the measures used are self-report, or proxy-reported by champions, and both responses concerning the organisation, the program, and the sending out of staff surveys all relied on the workplace champions. While necessary for the scalable online program, this does have significant potential for measurement error and bias. Importantly, staff perspectives were collected as well as the champion perspectives, and many staff findings were robust to missing data handling. Single-item or minimal item measures were chosen where possible in order to minimise participant burden in line with our preliminary work informing the trial.(21) One possibility for better gauging the impact of this program and others like it on productivity and health is to continue the broad, brief evaluation model but add optional evaluation tools with more detailed assessments. These may have measurement properties more suitable to detecting changes.

## Conclusion

Findings from this Australian national implementation trial found that the workplace champion-delivered BeUpstanding program was effective in supporting teams of desk-based workers to sit less and move more and in creating a supportive culture for this change. It also demonstrated that the program can be taken up and implemented by a wide range of users from across Australia and across industries. Learnings from the trial are being used to inform the next iteration of the program (BeUpstanding 2.0), which in turn will inform Phase 5 (sustainability) of the research-to-practice process.(12) This optimisation, which will be described in detail elsewhere, is designed to enhance scalability, inclusivity and the user experience.

## Data Availability

The datasets generated and analysed during the current study are available from the corresponding author on reasonable request.

## Declarations

### Ethics approval and consent to participate

The study was conducted according to the guidelines of the Declaration of Helsinki, and approved by the Human Research Ethics Committee of The University of Queensland (protocol code #2016001743 on the 9th January 2017). All participants provided informed consent. No individual or personally identifiable information is reported.

### Consent for publication

Not applicable

### Authors contributions

GNH, ADG, AA, DWD, EGE, NDG, LG, ADL, MM, NO, TS, LS: conception, funding, recruitment, interpretation.

LS: recruitment

ADG; SKM, JJ, LU: recruitment; data collection; project management; partner engagement

EAHW: data management and data analysis

GNH and EAHW wrote the paper with input from all authors.

## Acknowledgements

We acknowledge and thank the partner investigators and their associated support staff, academic investigators, project staff, and external consultants for their contribution to the BeUpstanding program of research. We also acknowledge and thank all the participants who have taken part in the program, particularly our workplace champions.

